# Dramatic reduction of psychiatric emergency consultations during lockdown linked to COVID-19 in Paris and suburbs

**DOI:** 10.1101/2020.05.19.20095901

**Authors:** Baptiste Pignon, Raphaël Gourevitch, Sarah Tebeka, Caroline Dubertret, Hélène Cardot, Valérie Dauriac-Le Masson, Anne-Kristelle Trebalag, David Barruel, Liova Yon, François Hemery, Marie Loric, Corentin Rabu, Antoine Pelissolo, Marion Leboyer, Franck Schürhoff, Alexandra Pham-Scottez

## Abstract

**Aims:** The COVID-19 pandemic and associated lockdown may have psychiatric consequences and increase the number of psychiatric emergency consultations. This study aimed to compare the number and characteristics of emergency psychiatric consultations during the four first weeks of the lockdown in three psychiatric emergency services from Paris and its suburbs, and to compare them to the same period in 2019.

**Methods:** Three psychiatric centers in Paris and its suburbs took part in the study. We compared the number of total psychiatric emergency consultations during the 4 first weeks of the lockdown in France to the corresponding 4 weeks in 2019. We also compared the number of consultations during these 4-week time periods in 2020 and 2019 across different diagnostic categories.

**Results:** In the 4 first weeks of the lockdown in France, 553 emergency psychiatry consultations were carried out, compared to 1224 consultations during the corresponding period of 2019, representing a 54.8 % decrease. This decrease was evident across all psychiatric disorders, including anxiety (number of consultations in 2020 representing 36.1 % of 2019), mood (41.1 %), and psychotic disorders (57.3 %). The number of suicide attempts also decreased (number of suicide attempts in 2020 representing 42.6 % of 2019). In comparison to 2019, the proportion of total consultations for anxiety disorders also decreased (16.6 % vs. 20.8 %), whilst the proportion of total consultations increased for psychotic disorders (31.1 % vs. 24.1 %).

**Conclusions:** The total number of psychiatric emergency consultations during lockdown dramatically decreased. The psychological consequences of lockdown may be delayed, indicating that psychiatric services should be prepared for a secondary increase in emergency presentations.

## INTRODUCTION

On March 17^th^, 2020 a national lockdown began in France in response to the COVID-19 pandemic. The virus underpinning the COVID-19 pandemic first appeared in China in November 2019, being declared as pandemic by WHO on March 11^th^ 2020 (Zhou et al., 2020). To date, France and Western Europe, per head population, are the most affected areas (Yuan et al., 2020).

A number of psychiatric consequences arising from the pandemic and lockdown have been proposed (Fagiolini et al., 2020; Fiorillo and Gorwood, 2020; Xiang et al., 2020). First, loneliness and social isolation caused by social distancing are long-established major risk factors for a number of psychiatric disorders, including anxiety and depression (Beutel et al., 2017; Courtet et al., 2020; Erzen and Çikrikci, 2018; Michalska da Rocha et al., 2018). Social isolation not only disrupts regular social rhythms, but can aggravate the negative symptoms evident in psychosis, including social withdrawal, apathy, and lack of social interest. The economic impact of the COVID-19 crisis may also increase psychiatric vulnerability (Pfefferbaum and North, 2020; Wickham et al., 2014). Confinement can also increase family/partner conflicts and violence. Quarantine and lockdown have other psychological consequences, such as boredom, anger, frustration, irritability, and sleep dysregulation, which are all associated with poorer psychiatric outcomes, including first episode emergence of psychiatric disorders as well as the exacerbation of pre-existing psychiatric conditions (Brooks et al., 2020; Rajkumar, 2020; Rolland et al., 2020). Contamination fear has additional stress associations, especially for health anxiety associated with anxious and obsessional symptoms, as well as some delusional symptoms (Brown et al., 2020; Fiorillo and Gorwood, 2020).

In addition to these stressors, psychiatric services have had to be extensively reorganized in response to the COVID-19 pandemic (Arango, 2020; Corruble, 2020; Fagiolini et al., 2020; Fiorillo and Gorwood, 2020; Freeman, 2020; Xiang et al., 2020), including in France (Chevance et al., 2020). A number of organizational changes have had to occur in order to maintain the continuity of public psychiatric care, including restricting consultations to severe cases and re-organization of health care via teleconsultation, as well as early hospital release and restrictions on new hospitalizations. Moreover, several daily care facilities, including psychiatric day hospital services and day-therapy day programs, have been closed to reduce contacts among patients, and between patients and mental health care professionals. Most private psychiatric consultations have been closed or re-organized via teleconsultation. Consequently, patients may experience difficulties in accessing psychiatric services, or worry about being fined for non-compliance of lockdown rules. Overall, such factors may create a treatment gap and/or lead to break in follow-up and ongoing treatment, thereby increasing emergency consultations (Font et al., 2018; Reger et al., 2020).

This study aimed to compare the number and characteristics of emergency psychiatric consultations during the four first weeks of the lockdown in three psychiatric emergency services from Paris and its suburbs, and to compare them to the same period in 2019.

## METHODS

### Study design

Three psychiatric emergency centers took part in the study: one in Paris, and two in adjacent suburban cities, Colombes (Northwest Paris) and Créteil (Southeast Paris). The Paris center is called CPOA (“*Centre Psychiatrique d’Orientation et d’Accueil****”***). It is located in Sainte-Anne hospital, and is the biggest emergency psychiatric units in Paris and its suburbs. The two suburban emergency centers are part of two University-affiliated hospitals of the Assistance Publique-Hôpitaux de Paris (Louis-Mourier for Colombes, and Henri-Mondor for Créteil). The Colombes center is the only center of the three to admit children.

### Data collection: sociodemographic and clinical characteristics

The data of this study was extracted anonymously from hospital registers. We assessed the number of emergency consultations during the 4 first weeks of the French lockdown, *viz* from Tuesday 17^th^ March to Monday 13^th^ April 2020 inclusive, and of the corresponding weeks of 2019, *viz* from Tuesday 19^th^ March to Monday 15^th^ April 2019). Age, gender, and provenance (i.e., the patient’s origin, such as patient’s home, public roads, etc.) were extracted for all patients visiting the emergency services. Patient’s provenance was only available for the Paris and Créteil centers.

We also extracted data concerning the presence of a recent (< 1 week) suicide attempt (except for Créteil center, as this data was not available) and psychiatric diagnosis of each patient. Psychiatric diagnoses utilized ICD-10 classification and were pooled as follows: psychotic disorders *(F20* to *F29)*, mood disorders *(F30* to *F39)*, anxiety and stress-related disorders *(F40* to *F48)*, personality disorders *(F60* to *F69)*, addictive disorders *(F10* to *F19)* and other.

Patients’ outcomes following the emergency consultations were also noted, including the rates of hospitalization, and as to whether this was with or without the patient’s consent (except for Créteil center, as this data was not available). For the Paris center, consultations were also rated as to whether this was a first psychiatric consultation or not.

### Ethical procedures

The study was performed in accordance with the Declaration of Helsinki. The data was extracted anonymously from registers, in accordance with the ethical and security standards of the French National Data Protection Authority. According to the INSERM ethics committee, this study does not need an opinion of a research ethics committee according to the French law.

### Statistical analyses

The sociodemographic, clinical, and outcome characteristics were compared using chi-square tests. For each variable, 2019 and 2020 rates of each category were compared.

## RESULTS

### Number of psychiatric emergency consultations

During the four first weeks of the national COVID-19 related lockdown, 553 emergency psychiatric consultations were carried out, representing less than half (45.2 %) of the corresponding weeks in 2019 (1224 consultations). This decrease was evident in each of the three centers, with the number of the consultations in 2020, compared to 2019, representing 38.8 % in Paris, 52.4 % in Créteil, and 63 % in Colombes.

Sociodemographic characteristics (i.e., sex-ratio and age-bands proportions) of subjects between the year 2019 and the year 2020 were not significantly different, except for the proportion of 16-25 years-old patients, which was lower in 2020 (21.7 % in 2020 vs. 27.5 % in 2019, p-value = 0.012, see **Table 1** for details).

**Table 1.**
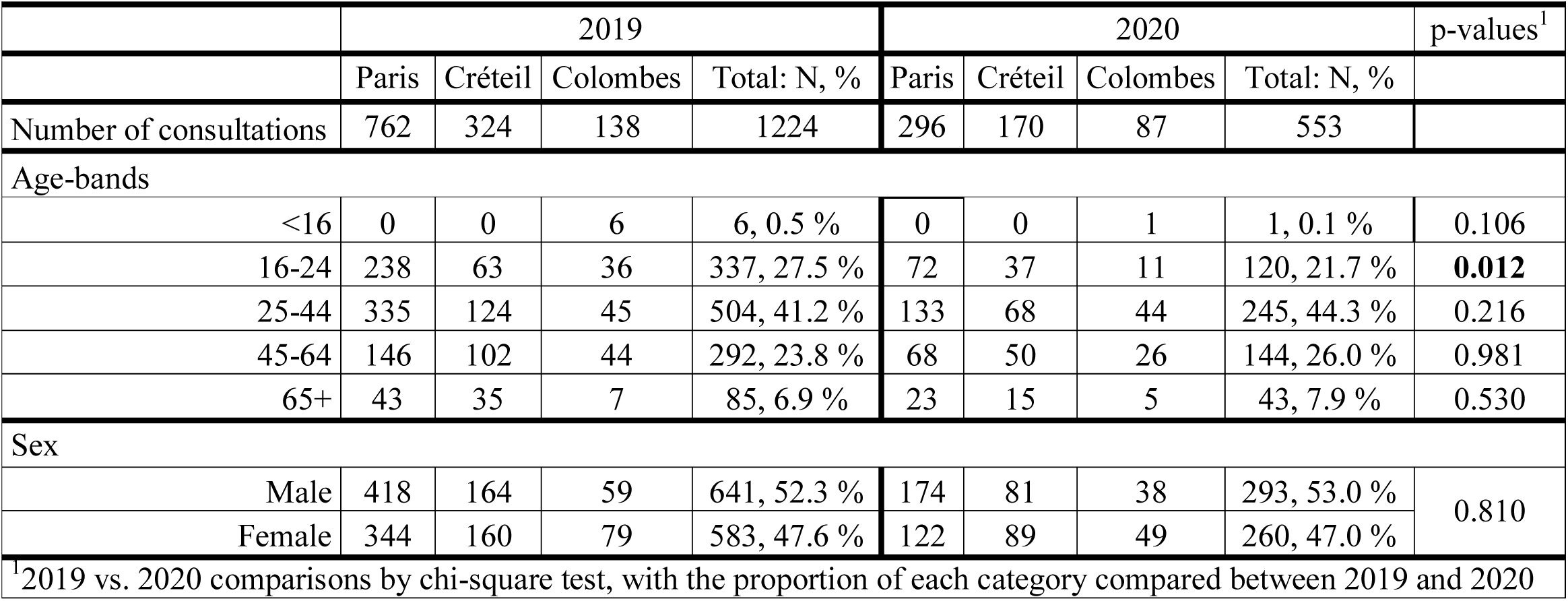
Number of emergency psychiatric consultations during the 4 first weeks of lockdown (2020), compared to the corresponding period in 2019.

### Clinical characteristics

Compared with 2019, the number of consultations for all the psychiatric diagnoses also decreased in 2020, especially for anxiety disorders (number of consultations in 2020 representing 36.1 % of consultations in 2019), mood disorders (41.1 %), and psychotic disorders (67.2 %). The number of consultations for each diagnostic category are represented in **Figure 1**. Total suicide attempts also decreased in 2020 to 42.6 % of those in 2019.

**Figure 1:**
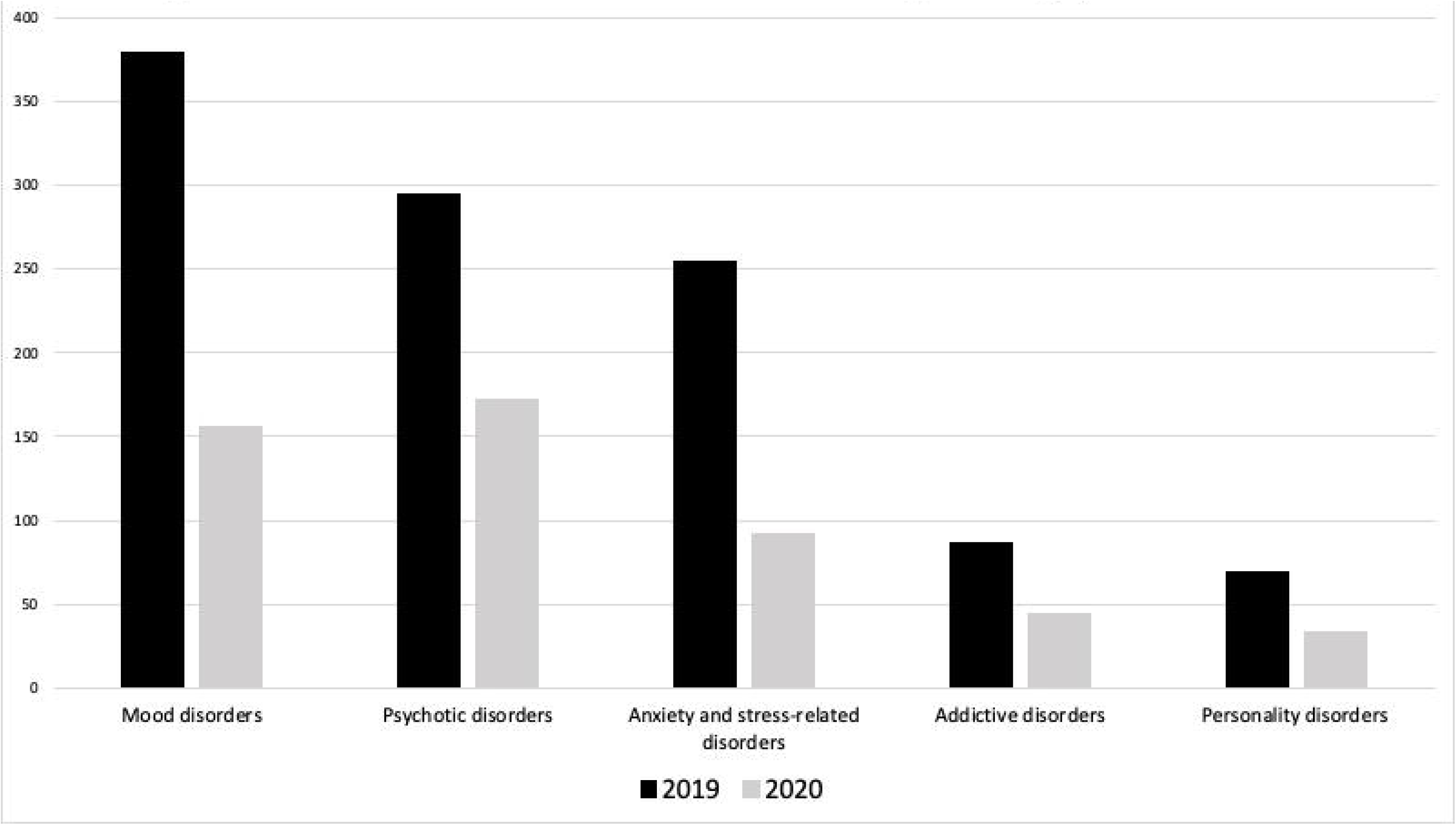
Bar chart of the number of consultations for each diagnosis category in 2019 and 2020350.

The diagnostic pattern of presentations changed in 2020, compared to 2019, with the percentage of consultations for psychotic disorders increasing (31.1 % in 2020 vs. 24.1 % in 2019, p-value = 0.002), in contrast to the decreases evident for anxiety and stress-related disorders (16.6 % vs. 20.8 %, p-value = 0.039). The rate of first-episode psychiatric consultations also significantly decreased in 2020 (13.8 % vs. 20.1 %, p-value = 0.018). Details concerning clinical and outcome characteristics are shown in **Table 2**.

**Table 2.** Clinical characteristics and orientation of patients having a psychiatric consultation in an emergency service.

|  |  |  |  |  |  |  |  |  |  |
| --- | --- | --- | --- | --- | --- | --- | --- | --- | --- |
|  | 2019 |  |  |  | 2020 |  |  |  | p-values<sup>1</sup> |
|  | Paris | Créteil | Colombes | Total: N, % | Paris | Créteil | Colombes | Total: N, % |  |
| Diagnoses |  |  |  |  |  |  |  |  |  |
| Mood disorders | 230 | 112 | 37 | 379, 31.0 % | 87 | 49 | 20 | 156, 28.2 % | 0.241 |
| Psychotic disorders | 184 | 69 | 42 | 295, 24.1 % | 98 | 40 | 34 | 172, 31.1 % | **0.002** |
| Anxiety and stress-related disorders | 175 | 48 | 32 | 255, 20.8 % | 52 | 29 | 11 | 92, 16.6 % | **0.038** |
| Addictive disorders | 54 | 24 | 9 | 87, 7.1 % | 15 | 16 | 13 | 44, 8.0 % | 0.402 |
| Personality disorders | 50 | 12 | 7 | 69, 5.6 % | 21 | 6 | 6 | 33, 6.0 % | 0.077 |
| Other | 52 | 22 | 7 | 81. 6.6 % | 20 | 11 | 0 | 31, 5.6 % | 0.283 |
| Unavailable data | 17 | 37 | 4 | 58, 4.7 | 3 | 19 | 3 | 25, 4.5 % | 0.840 |
| Hospitalization |  |  |  |  |  |  |  |  |  |
| Yes | 360 | 144 | 58 | 562, 45.9 % | 121 | 99 | 45 | 265, 47.9 % | 0.872 |
| No | 329 | 139 | 80 | 548, 44.8 % | 153 | 59 | 42 | 254, 45.9 % |  |
| Unavailable data | 73 | 41 | 0 | 114, 9.3 % | 22 | 12 | 0 | 34, 6.1 % |  |
| Hospital admission without consent<sup>2</sup> |  |  |  |  |  |  |  |  |  |
| Yes | 158 | NA | 25 | 183, 43.8 % | 65 | NA | 25 | 90, 54.2 % | **0.022** |
| No | 202 |  | 33 | 235, 56.2 % | 56 |  | 20 | 76, 45.8 % |  |
| Suicide attempts |  |  |  |  |  |  |  |  |  |
| Yes | 53 | NA | 22 | 75, 8.4 % | 23 | NA | 9 | 32, 8.4 % | 0.812 |
| No | 651 |  | 114 | 765, 85.2 % | 266 |  | 78 | 344, 89.8 % |  |
| Unavailable data | 58 |  | 0 | 58, 6.5 % | 7 |  | 0 | 7, 1.8 % |  |
| Provenance |  |  |  |  |  |  |  |  |  |
| Home | 429 | 195 | NA | 624, 57.5 % | 203 | 101 | NA | 304, 65.2 % | **0.004** |
| Other hospital | 133 | 11 | NA | 144, 13.3 % | 21 | 5 | NA | 26, 5.8 % | <0.001 |
| Institution | 39 | 3 | NA | 42, 3.9 % | 11 | 0 | NA | 11, 2.4 % | 0.134 |
| Public road | 120 | 13 | NA | 133, 12.2 % | 55 | 3 | NA | 58, 12.4 % | 0.912 |
| Other | 41 | 102 | NA | 143, 13.2 % | 6 | 61 | NA | 67, 12.1% | 0.523 |
| First psychiatric consultation rate |  |  |  |  |  |  |  |  |  |
| Yes | 153 | NA | NA | 153, 20.1 % | 41 | NA | NA | 41, 13.9 % | **0.018** |
| No | 600 | NA | NA | 600, 78.7 % | 252 | NA | NA | 252, 85.1 % |  |
| Unavailable data | 9 |  |  | 9, 1.2 % | 3 |  |  | 3, 1.0 % |  |
| Presence of an accompanying person |  |  |  |  |  |  |  |  |  |
| Yes | 372 | NA | NA | 372, 48.8 % | 126 | NA | NA | 126, 42.6 % | 0.064 |
| No | 390 | NA |  | 390, 51.2 % | 170 |  |  | 170, 57.4 % |  |
| Legends: <sup>1</sup>2019 vs. 2020 comparisons by chi-square test, with the proportion of each category compared between 2019 and 2020; <sup>2</sup>among hospitalizations. |  |  |  |  |  |  |  |  |  |

### Orientation decisions after emergency consultations

The rate of hospital admission after emergency consultation was not significantly different between 2019 and 2020. However, in 2020, hospitalization without patients’ consent significantly increased (54.2 % in 2020 vs. 43.8 % in 2019, p-value = 0.023).

## DISCUSSION

Given the multi-faceted stressors associated with lockdown, the above results show a surprising, and dramatic, 54.8 % drop in the number of psychiatric emergency consultations, during the first 4 weeks of the COVID-19 pandemic, compared to the same period in 2019. As indicated by the presented data, this decrease is evident in the 3 emergency departments of Paris and its suburbs, covering both the psychiatric emergency service and general hospital emergency services of these units. Further, this decrease is evident across all psychiatric diagnostic categories, and concerns also suicide attempts. The percentage of anxiety disorders was lower in 2020 than in 2019, whilst the percentage of patients consulting for psychotic disorders was higher in 2020 than in 2019, as was the rate of hospitalization without consent.

Data from other countries indicates that this decrease is not specific to psychiatry. In the West China Hospital emergency department, a greater than 50 % decrease in daily total consultations was reported, coupled to an elevation in consultations for fever and/or COVID-19 symptoms (Cao et al., 2020). A similar phenomenon has been observed in England, where lockdown led to a 25 % in general emergency consultations during the week (Thornton, 2020). Clearly, a fear of contamination in emergency departments has contributed to this. A huge rise in psychiatric emergency consultations may be expected after lockdown, and perhaps in the later phases of lockdown. Moreover, the number of consultations, which are not strictly medical emergencies, may also have decreased. In France, as in many other countries, recent decades have seen a significant increase in the number of emergency department consultations (Derlet and Richards, 2000; Hoot and Aronsky, 2008). This increase is contributed to by multiple complex factors, including a deterioration in accessibility of primary care services, leading to unnecessary hospital emergency departments visits (Cunningham et al., 1995). The treatment gap in psychiatry, the gap between experiencing a psychiatric disorder and using treatment services for this disorder, has been extensively described in France and elsewhere (Font et al., 2018; Kohn et al., 2004). The results in the current study seem in line with this, given the significant increase of the proportion of consultations for psychotic disorders and of hospitalizations without consent, coupled to the significant decrease in primary psychiatric consultations in the largest center of our study (Paris). For the most severe psychiatric disorders, the emergency consultations are more necessary, and the number of consultations is less attenuated.

The development of telemedicine would also seem to have contributed to our results. Psychiatric services have been quickly mobilized to provide telemedicine consultations, which seems well accepted by patients and mental health care professionals, with the present results possibly indicative of the greater utilization of telemedicine consultations during the COVID-19 pandemic (Kavoor et al., 2020). For depression or anxiety disorders, telemedicine consultations could even have more efficacy than traditional consultations (Ekeland et al., 2010; Fortney et al., 2013). This viability and feasibility of telemedicine consultations are likely to emerge subsequent to the COVID-19 triggered lockdown, possibly indicating a role for their sustained implementation. This is a change that may emerge from the COVID-19 pandemic and it will be interesting to determine its impact on factors such as the rates of patients lost to follow-up.

As some people may find new strengths and coping strategies during disasters (Pfefferbaum and North, 2020), the current results may arise from an elevation in resilience capacity. This was observed in New York following the September 11^th^ terrorist attack, where the expected surge in psychiatric presentations, including post-traumatic stress symptoms and/or disorders, did not emerge (Bonanno et al., 2006). Such psychological resilience is described as having both individual and collective aspects (Williams and Drury, 2009).

Overall, despite the expectation of lockdown-induced stress increasing relapse risk across psychiatric conditions, the numbers of patients seeking emergency psychiatric consultations have decreased during lockdown. This is important to document, as will be the amount of consultation in the post-pandemic period. The reasons underpinning this dramatic reduction, such as telemedicine efficiency and case-management strategies, may be incorporated to improve quality and organization of health care provision. However, the psychological consequences of lockdown may occur later, where a secondary increase of emergency psychiatric presentations may occur. Clearly, COVID-19 has had an impact on psychiatric service utilization and will continue to do so (Chevance et al., 2020), whilst also having possible implications for the nature of psychiatric service organization.

## Data Availability

N/A

## Acknowledgements

We want to thank Dr. Yohan Dabi for his advice, and Dr. George Anderson for his prompt editing work.

## Authors contributions

- Conception and design of the study: Baptiste Pignon, Raphaël Gourevitch, Franck Schürhoff and Alexandra Pham;
- Extraction of the data: Sarah Tebeka, Hélène Cardot, François Hemery, Marie Loric, Valérie Dauriac-Le Masson, David Barruel;
- Statistical analyses: Baptiste Pignon;
- First draft of the manuscript: Baptiste Pignon and Franck Schürhoff;
- Writing and revision of the paper: all authors.

## Availability of Data and Materials

The data is available on request.

